# Identification of new angio-architectural features of at-risk cranial dural arteriovenous fistulas using machine learning approaches

**DOI:** 10.1101/2023.08.06.23293535

**Authors:** Katharina Frank, Paiman Shalchian-Tehran, Mihai Manu, Zafer Cinibulak, Jörg Poggenborg, Makoto Nakamura

**Author notes:** These authors contributed equally (Katharina Frank, Paiman Shalchian-Tehran, Mihai Manu).

## Abstract

**Background:** Cranial dural arteriovenous fistulas (dAVF’s) are rare complex vascular malformations that have a bleeding risk with potential lethal consequences. Despite this, the vascular architectural features associated with the rupture risk are not always clearly defined.

**Methods:** We retrospectively analyzed cranial arteriovenous fistulas in terms of their anatomical and angio-architectural features as evaluated on conventional subtraction angiography: Location of the fistula, fistula architecture, venous ectasia, reflux in cortical draining veins, presence of pial feeders, outflow stenosis, presence of a major sinus thrombosis, flow-associated arterial aneurysms as well as presenting symptoms. Patterns in the data were identified after multiple components analysis followed by automatic k-means clustering and their predictive power was confirmed using a neural network and a random forest classifier.

**Results:** New relevant features predictive of hemorrhage (venous outflow stenosis and fistula architecture) were identified using distinct but surprisingly converging modeling paradigms. Both the neural network and the random forest classifier achieved a relatively high performance metric, with area under the receiver operating characteristic curve (ROC AUC)) of 0.875 [95% CI, 0.75-1.0]. The relevance of these findings was verified by performing a multiple correspondence analysis followed by k-means clustering in the angiographic feature vector space. There was good agreement between the ground truth (hemorrhage) and the cluster labels (adjusted Rand score 0.273, purity index 0.82).

**Conclusion:** Machine learning approaches confirmed the importance of previously described features (reflux in a cortical vein and venous ectasia) but also uncovered novel relevant characters (outflow stenosis and fistula architecture) for the hemorrhage risk of dAVF’s.

## Introduction

Cranial dural arteriovenous fistulas (dAVF’s) are abnormal direct arteriovenous shunts within the dura with multifactorial pathogenesis, which are acquired and can develop spontaneously, after traumatic head injury or e.g. after thrombosis of the venous sinus. dAVFs are making up around 10% of all intracranial vascular malformations and have an incidence around 0,15-0,29 per 100,000 adults per year(1, 2). Patients harboring these malformations are typically middle-aged, but the lesions can, although less frequently, occur in children. The symptoms are presumed to depend on the presence or absence of venous hypertension and can vary from benign symptoms (e.g. tinnitus) up to severe hemorrhage in any of the three compartments (intracerebral, subdural and/or subarachnoidal) or non-hemorrhagic neurologic deficits (NHNDs) such as seizures(2). Regardless of presentation, the first line imaging tools are computed tomography and CT-angiography, less frequently magnetic resonance imaging/angiography/venography (MRI, MRA, MRV), whereas digital subtraction angiography (DSA) represents the gold standard for diagnosis and therapy(3).

There are several classification and grading systems, aiming to stratify the risk and guide treatment decisions, of which the Cognard and the Borden classifications are commonly used(4, 5). Higher-grade fistulas, associated with venous hypertension and consequently ectatic cortical veins, are presumed to carry an increased hemorrhage risk, have poorer outcomes and thus are prone to a more aggressive treatment approach(6-8).

We hypothesized that venous hypertension is the result of nonlinear interactions of features that have a direct angiographic correspondence (not necessarily in the form of dilated cortical veins, as evidenced by symptomatic fistulas that do not exhibit this character), that the hemorrhage risk of dAVF’s may largely depend on these features, and that these dependencies, given their nonlinear nature, are not likely captured by traditional statistical methods. Our goal was to explore the use of machine learning in identifying potential relevant, hidden, angio-architectural features of dural arteriovenous fistulas helping neurosurgeons and interventional neuroradiologists in clinical decision-making process.

## Methods

Our retrospective cohort comprised 23 cranial dural arteriovenous fistulas that underwent digital subtraction angiography at the Department of Neurosurgery of the Cologne-Merheim Medical Center between January 2017 and December 2022. Additionally, all patients that presented with hemorrhage or non-hemorrhagic neurological deficits received at least a CT scan or CT-angiography. The recorded imaging parameters were location of the fistula, fistula architecture, venous ectasia, cortical venous drainage, presence of pial feeders, outflow stenosis, presence of a major sinus thrombosis and flow-associated arterial aneurysms. The clinical variables included age, sex and symptoms at presentation.

The machine learning was implemented by means of custom-written routines in Python. We applied a dimensionality reduction algorithm (multiple correspondence analysis (MCA), prince python package (9)) to identify the direction of largest variance by performing a singular value decomposition of the chi-square distance matrix followed by automatic k-means clustering in the resulting low-dimensional space.

Both the random forest classifier as well as the neural network (based on the multi-layer perceptron (MLP) classifier, supplemental figure 1 A) were implemented by means of the scikit-learn Python package(10). The shallow feedforward neural network had one output and one input layer as well one hidden layer comprising 12 neurons with rectified linear activation functions (ReLU) and standard backpropagation with gradient descent (Adam) with the goal to minimize the objective function (binary cross-entropy loss).

## Results

The median age at the time of diagnosis was 59 years (range: 28-84). There were 70% males (16/23) and 30% female (7/23) patients. The angiographic features, anatomical location, Cognard class of the fistula as well as the presenting symptoms are summarized in Table 1 and illustrated in Figure 1.

**Table 1.** Summary of the anatomical and angiographic characters of the dAVF. (*)Venous ectasia was defined as a cortical dilated vein of more than 5mm diameter. (**)Outflow stenosis was considered where there was a 50% abrupt change in the diameter of the cortical draining vein. (***) Total bleeding events is less than the sum of compartment events (parenchyma, subdural and/or subarrachnoidal space) due to concomitant presence of blood in more than one compartment.

| Pattern | Number (%) |
| --- | --- |
| Fistula location<br>• transverse-sigmoid junction<br>• ethmoid<br>• tentorial<br>• petrosal<br>• convexity cortical vein<br>• foramen magnum | 6/23 (26%)<br>1/23 (6%)<br>7/23 (30%)<br>2/23 (8%)<br>5/23 (21%)<br>2/23 (8%) |
| Fistula architecture<br>• multi feeder spread-out<br>• multi feeder converging<br>• single feeder | 14/23 (60%)<br>8/23 (36%)<br>1/23 (4%) |
| Reflux in a cortical vein | 20/23 (86%) |
| Venous ectasia* | 12/23 (52%) |
| Pial feeder | 9/23 (39%) |
| Venous outflow stenosis** | 15/23 (65%) |
| Venous sinus thrombosis | 6/23 (26%) |
| Flow-associated aneurysm | 3/23 (13%) |
| **Presenting symptoms** |  |
| Hemorrhage***<br>• parenchymal<br>• subdural<br>• subarachnoidal | 9/23 (39%)<br>5/23 (22%)<br>7/23 (30%)<br>2/23 (8%) |
| Seizures | 3/23 (13%) |
| Tinnitus | 8/23 (34%) |
| Visual field defects | 1/23 (4%) |
| Asymptomatic | 2/23 (8%) |
| **Cognard classification** |  |
| • I<br>• IIa<br>• IIb<br>• III<br>• IV<br>• V | 1/23 (4%)<br>1/23 (4%)<br>5/23 (21.7%)<br>4/23 (17%)<br>11/23 (48%)<br>1/23 (4%) |

**Figure 1.**
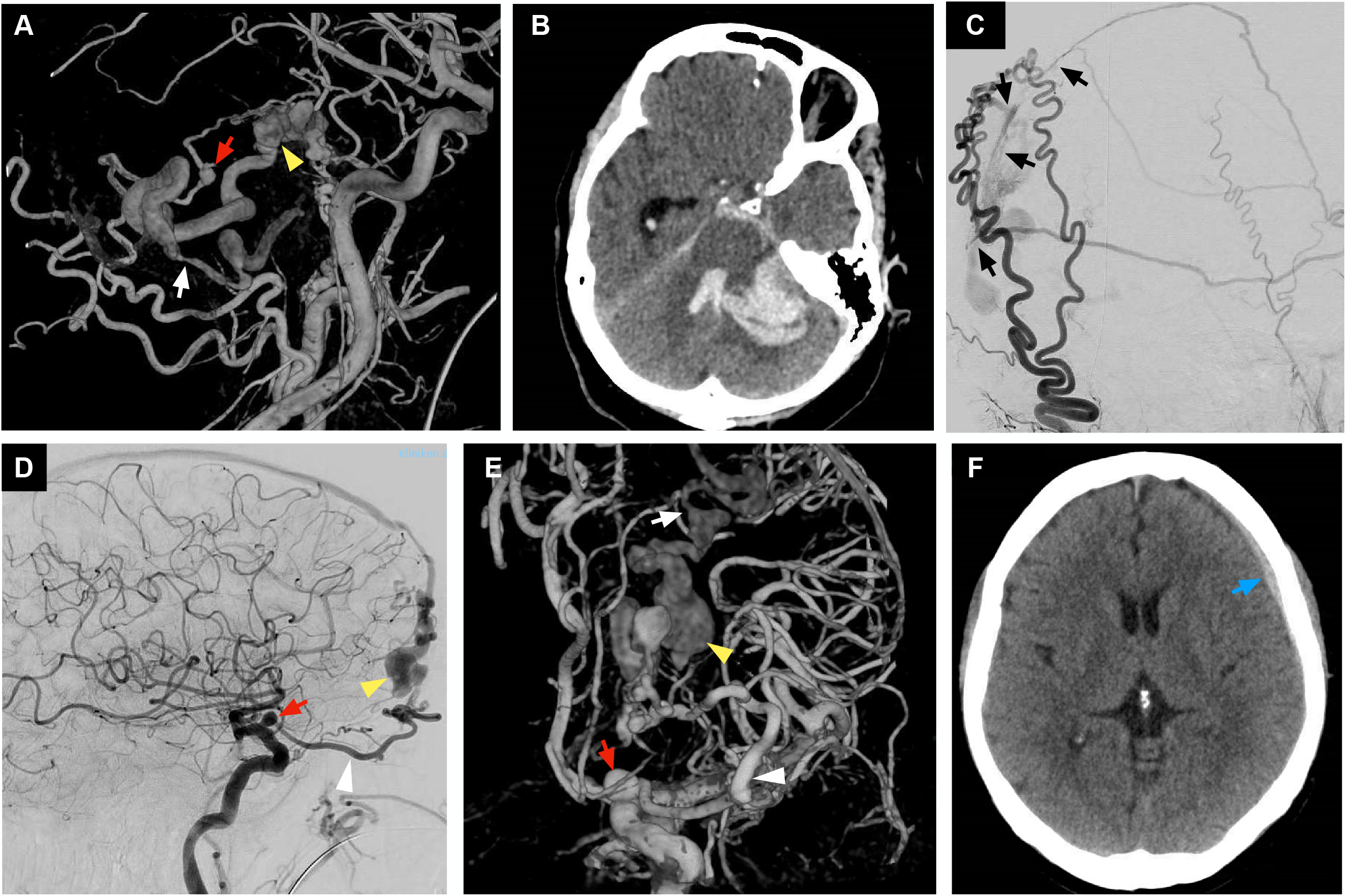
Angiographic characters of dural arteriovenous fistulas. (A) 3D reconstruction of a common carotid injection showing ectatic cortical vein (yellow arrowhead), segmental stenosis of the draining vein (white arrow) as well as a flow-associated aneurysm (red arrow). (B) Same patient as in A, CT scan showing cerebellar hemorrhage. (C) External carotid injection in another case showing a multiple feeder (black arrows) spread out type of fistula. (D) Left internal carotid injection showing a flow-associated aneurysm as well as dilated cortical vein (color-coding as in A). Note the prominent ophthalmic artery (white arrowhead). (E) 3D reconstruction of the previous angiography, viewed anterograde, demonstrating additionally a segmental stenosis of the draining vein (white arrow, color-coding as in A). (F) Same patient as in D and E, CT scan at presentation demonstrating a small left sided acute, non-traumatic subdural hematoma.

In order to extract hidden and relevant features for the hemorrhage risk from the conventional subtraction angiography, we trained (80% of the data) and tested (20% of the data) a shallow feed-forward neural network (ANN), where the input was a list of fistula characters and the output was the probability of being in one of two categories (hemorrhage vs non-hemorrhage). The model achieved a relatively high receiver operating characteristic area under the curve (ROC AUC) score of 0.875 [95% CI, 0.75-1.0] and an accuracy of 0.8, indicating good performance in predicting the outcome of interest. Independently checking the model performance with 5-fold cross-validation resulted in a cross-validation accuracy of 0.82 ± 0.094 (supplemental figure 1 B and C). We next looked at the importance scores of the different input features, as represented by the sum of the weight values for individual input features across all the layers. We found the largest score for reflux in a cortical draining vein, venous outflow stenosis and presence of pial feeders.

Next, we asked if the same results can be reproduced by applying a different supervised learning algorithm. To this end, we turned to a random forest classifier, applied to the same dataset. Again, the model achieved a relatively high receiver operating characteristic area under the curve (ROC AUC) score of 0.875 [95% CI, 0.75-1.0] indicating also good performance in predicting the outcome. Interestingly and importantly, the most significant three features in making accurate prediction within the model were venous outflow stenosis, venous ectasia and presence of pial feeders.

Thus, the analysis of feature importance of two distinct models revealed a consistent pattern: presence of venous outflow stenosis and pial feeders, next to venous ectasia and reflux in a cortical draining vein emerged as pivotal contributors to the classification task.

To verify these findings, we performed a dimensionality reduction by means of multiple correspondence analysis followed by k-means clustering in the dAVF’s feature vector space. We found that the characters of the fistula that correlate best with the first two components (the eigenvectors with the largest eigenvalue, thus explaining most of the variance in the data) were the architecture of the fistula, venous outflow stenosis, venous ectasia and reflux in a cortical draining vein (supplemental figure 2 A). The automatic k-means clustering in this low-dimensional space was verified by checking the agreement between the ground truth (actual outcome) and the cluster labels (adjusted Rand score of 0.273 (range -1 to 1, values closer to 1 indicate better fit), purity index=0.82) (supplemental figure 2 B).

## Discussion

Here we describe a novel approach to identify bleeding risk factors for cranial dural arteriovenous fistulas, by revealing patterns and their features in the data that best discriminate hemorrhagic/non-hemorrhagic lesions using MCA coupled with automatic k-means clustering and confirming the relevance of these hidden, important angiographic features that best predict hemorrhage using a shallow artificial neural network and a random forest classifier. This approach achieved 80% accuracy in correctly classifying those dAVF’s that present with hemorrhage, and showed the importance of new, previously ignored features such as venous outflow stenosis and angio-architecture of the fistula. Additionally, the machine learning approach confirmed the relevance of previously deemed important characters such as pial feeder and venous ectasia(4, 11, 12). Intriguingly, this analysis suggest, for the first time, that the same character (venous outflow stenosis) that is associated with increased rupture risk in other, more frequent shunting vascular malformation such as the cerebral arteriovenous malformation (AVM)(13) presumably play the same role in dAVF’s.

The presence of pial feeding arteries(14) and cortical venous drainage with or without abnormal venous dilatation(6) have been considered independent risk factors for predicting the natural history of dAVF’s. As such, some of these characters (reflux in a cortical vein, venous ectasia) are part of the widely used classifications of Borden and Cognard that guide treatment decision(5, 6). Despite the caveats of applying machine learning to small datasets (but see Olson et al., 2018 for a sound argument against this)(15), our work confirms and extends the findings of prior studies and makes the argument that the concomitant presence of these four features on the conventional angiography may be sufficient to achieved greater discrimant power regarding the bleeding risk. The congruence between the classification performance of the shallow neural network and random forest classifier underscores the stability of our findings and emphasizes their robust discriminatory power across distinct model paradigms. To our knowledge, this is the first study in literature that applies machine learning in evaluating features of dAVF’s predictive for hemorrhage risk and is an approach that can be applied to other lesions and organs.

## Conclusions

Using machine learning, we show the importance of two new angiographic features (venous outflow stenosis and fistula architecture) for the evaluation of dAVF’s. Their presence, independent or in addition to cortical venous drainage, venous ectasia and pial feeders, can predict with relative high accuracy the hemorrhagic risk of these lesions. This shows that simple machine learning algorithms can identify potential relevant, hidden, angio-architectural features of dural arteriovenous fistulas helping neurosurgeons and interventional neuroradiologists in the clinical decision-making process. Our approach can be applied to larger and more complex datasets as well to other vascular or non-vascular lesions.

## Data availability statement

Data is available upon reasonable request. The authors confirm that the data used in this study cannot be made available in the manuscript, the online supplemental files or in a public repository due to the German data protection law (“Bundesdatenschutzgesetz”, BDSG)

## Ethics approval

This study was approved by the Ethics Committee of the Witten/Herdecke University

