## Supplemental Figure 1 and Supplemental Figure 2 for "Identification of new angio-architectural features of at-risk cranial dural arteriovenous fistulas using machine learning approaches"

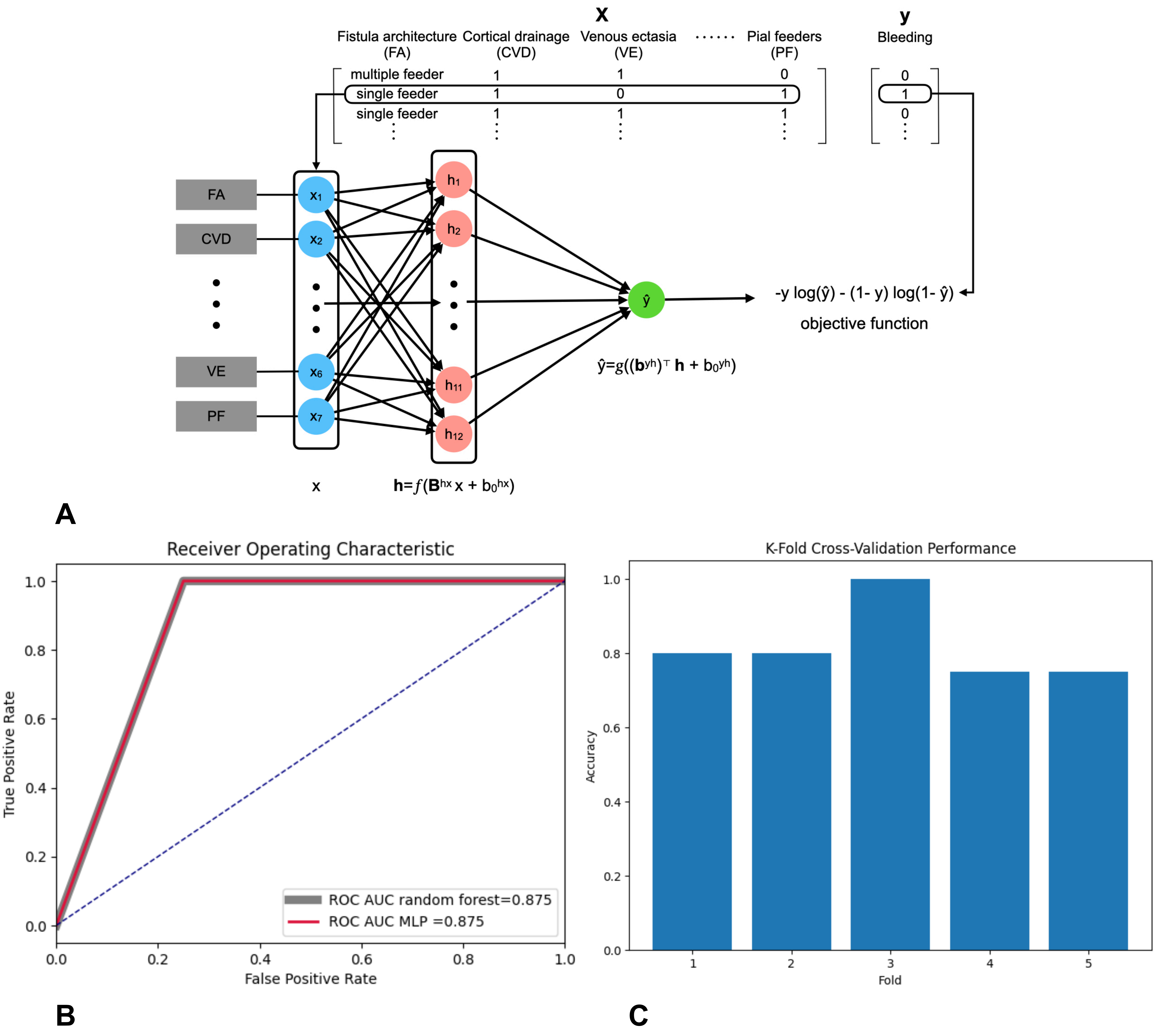


Supplemental figure 1. Machine learning classification and clustering of cranial dural arterio-venous fistulas. (A) Illustration of the shallow feed-forward neural network (multi-layer perceptron, MLP). The input layer (blue) takes a binary vector of the predictors (fistula characters, top) x, computes the h vector as a weighted sum of the input by multiplying with a weight matrix B and then passing the result through a ReLU function $f$. In the final stage, the output (green) is computed as a weighted sum of h passed through a sigmoid activation function $g$. The output value corresponds the probability of meeting the outcome criteria (bleeding, y). (B) Graphical representation of the random forest classifier (gray curve) and of the MLP classifier (red curve) performance as area under the curve of the ROC. (C) k-fold cross-validation performance showing how the MLP classifier model generalizes to different subsets of the data as well as its stability and reliability.

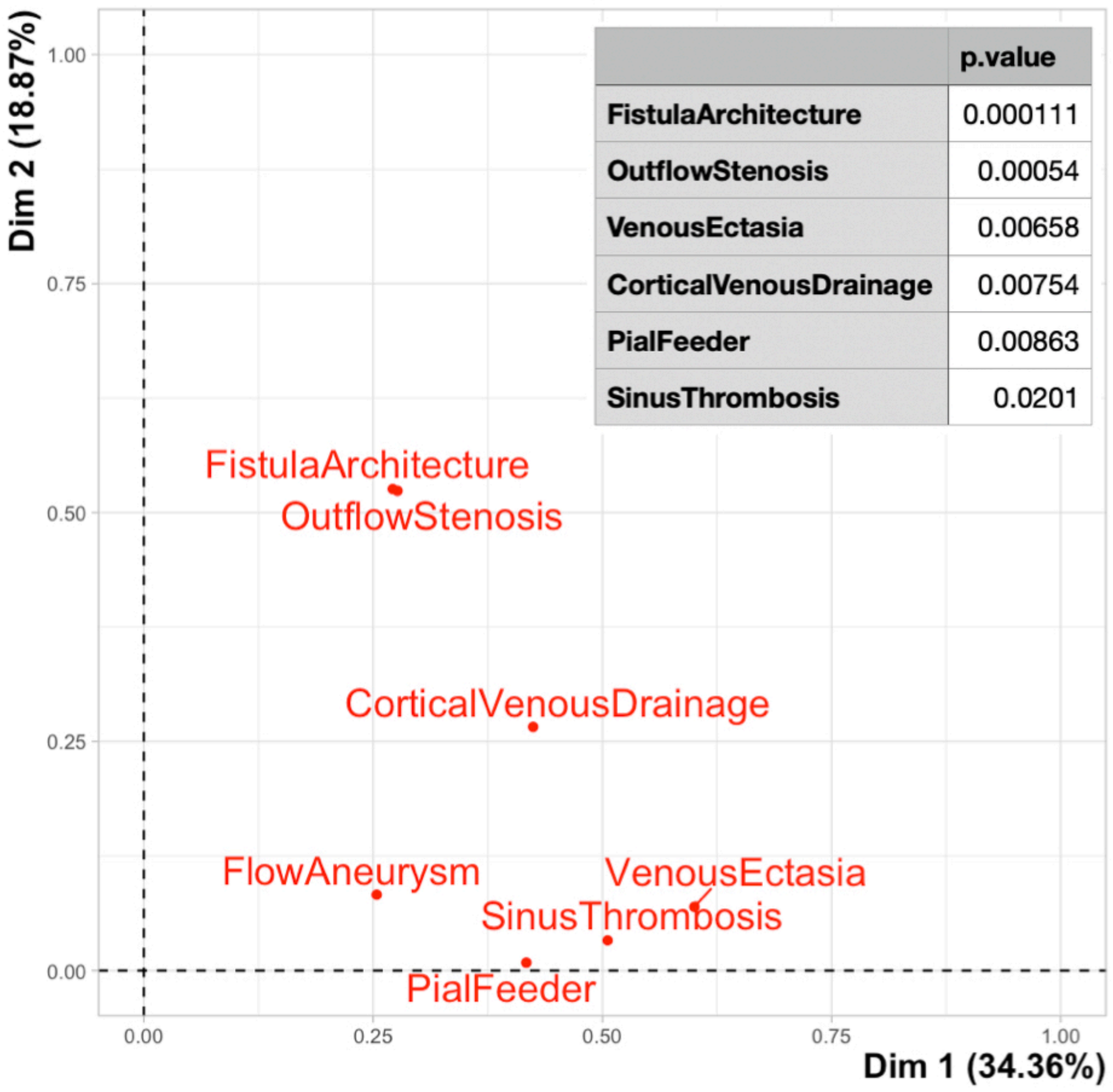


A

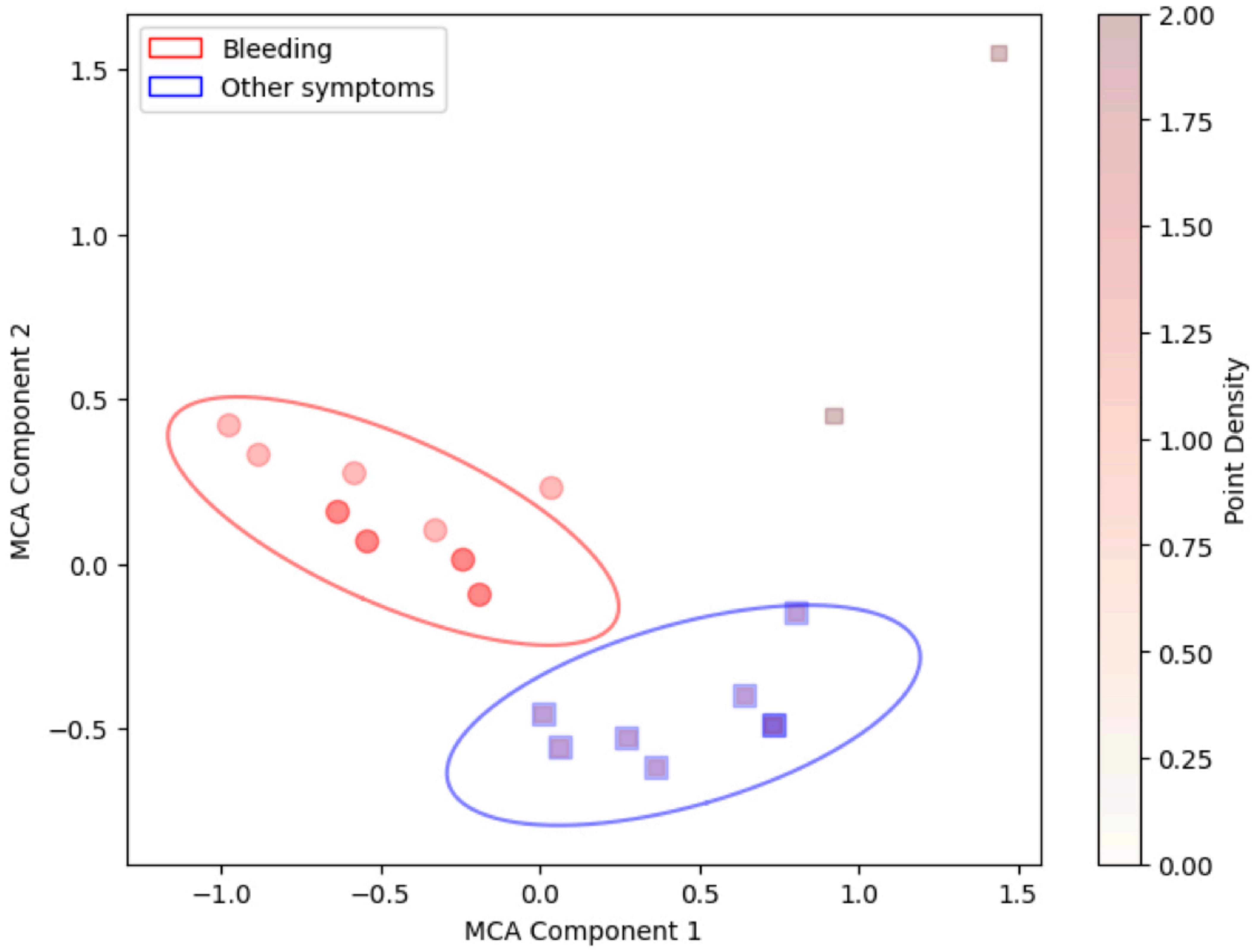


B

Supplemental figure 2. Machine learning classification and clustering of cranial dural arterio-venous fistulas. (A) Predictor variable representation after dimensionality reduction using multiple correspondence analysis (MCA) showing the correlation between the angiographic features and the principal dimensions. The individual coordinates are given by the squared correlations. Dim1 and 2 correspond to the first principal components with their percentage of explained variance. Insert shows the p-values (chi-squared test) of the features (significance of the relationship between categorical variables and dimensions in the MCA solution) (D) K-means clustering (MCA component 1 and 2 correspond to the first 2 principal components) in the multiple correspondence analysis space. Each oval indicates 1 standard deviation of a 2D gaussian fit. The 2 outlying points correspond to lesions without symptoms.
